# Abnormality Detection in Time-Series Bio-Signals using Kolmogorov-Arnold Networks (KANs) Based Models for Resource-Constrained Devices

**DOI:** 10.1101/2024.06.04.24308428

**Authors:** Zhaojing Huang, Jiashuo Cui, Leping Yu, Luis Fernando Herbozo Contreras, Girish Dwivedi, Omid Kavehei

## Abstract

This study investigates Kolmogorov–Arnold Networks (KANs) for biosignal analysis, using electrocardiogram signals as a case study. KANs provide flexibility and require few parameters, making them suitable for wearable and edge devices. A simple KAN model with one hidden layer of 64 neurons was trained on the TNMG dataset and tested on the CPSC 2018 dataset, achieving an F1-score of 0.75 and AUROC of 0.95 on TNMG, and an F1-score of 0.62 and AUROC of 0.84 on CPSC. The model also showed robustness to missing channels, maintaining reasonable performance with only a single ECG lead. Compared with traditional Multi-Layer Perceptrons (MLPs) and Neural Circuit Policies (NCPs), KANs demonstrated greater flexibility, adaptability, interpretability, and efficiency. Additionally, a shallow network (CKAN) that integrates a single Conv2dLSTM layer with a small set of KAN neurons, mirroring two architectures built with different NCP neurons for TinyML, achieved an F1-score of 0.84 and an AUROC of 0.97 on TNMG, and an F1-score of 0.72 and an AUROC of 0.92 on CPSC. Incorporating learnable sparsity, a key feature of NCP neurons, into KAN neurons surprisingly enhanced both performance and generalization. Even after pruning sparse weights, the model maintained strong performance, surpassing the counterpart without sparsity.

## 1 Introduction

There are many techniques for monitoring cardiac activity, with Electrocardiography (ECG) being the most widely used option due to its non-invasive and costeffective characteristics. The 12-lead ECG is the preferred method for evaluating the heart’s electrical activity in clinical settings [1]. Detecting and treating rhythm abnormalities is a key function of cardiac care units. Cardiac rhythm abnormalities are common in community-dwelling adults, affecting more than 2% of the population [2]. ECG signals can also help detect seizures [3]. The automatic computer analysis of standard 12 lead electrocardiogram is becoming increasingly important in the medical diagnosis process [4]. However, when morphological disturbances in the ECG signal become complex, interpretations can vary significantly among physicians [5]. This variability underscores the need for a system capable of analyzing ECG signals with high accuracy, reducing errors, and enabling timely detection and treatment of issues [6].

Multi-layer Perceptrons (MLPs) are fundamental to deep learning models and have been extensively utilized in applications like ECG signal analysis. Known for their capability to approximate nonlinear functions, MLPs are effective in detecting ECG abnormalities, which is crucial for cardiac diagnosis and monitoring [7, 8, 9]. However, MLPs often have limitations, such as fixed activation functions and less interpretable structures, which can hinder performance improvements and clinical applicability [10].

In response to these limitations, more compact and efficient neural network models such as Neural Circuit Policies (NCPs) have been developed. NCPs which is designed for energy-efficient computation, have demonstrated impressive performance in various tasks, including ECG signal analysis [11]. By integrating principles from neuroscience, NCPs achieve high accuracy with fewer parameters, rendering them suitable for deployment on resource-constrained devices [12]. Additionally, there is potential for merging NCPs with Spiking Neural Networks to enhance power efficiency [13].

Kolmogorov-Arnold Networks (KANs) offer a promising alternative. Inspired by the Kolmogorov-Arnold representation theorem, KANs use learnable activation functions on network edges rather than fixed node functions.

This design enhances flexibility, adaptability, and interpretability, which are critical in medical applications where understanding model decisions is essential. Recent studies, such as those by Liu et al. [14], have shown KANs outperforming traditional MLPs in tasks like data fitting and solving partial differential equations, suggesting their effectiveness in optimizing ECG analysis and improving cardiac diagnoses.

This work examines the performance of KANs, comparing them with MLPs and NCPs in the context of ECG signal analysis, focusing on detecting cardiac rhythm abnormalities. These comparisons are graphically depicted in Fig. 1 and detailed in Table 1. KANs hold promise for future hardware implementation due to their potential for reduced parameters and efficiency. Their adaptability and efficiency are important features and, hence, candidates for deployment in hardware, providing practical solutions for real-time monitoring and diagnosis in clinical environments. Evaluating the architectures under diverse conditions aims to assess the potential of KANs to enhance ECG analysis in cardiac healthcare. Given the increasing demand for efficient and real-time monitoring solutions, this work envisions the KANs deployment in wearable edge-devices. These devices are designed to swiftly detect anomalies and effectively identify cardiac abnormalities, leveraging KAN’s compact architecture and reduced computational complexity. This work also integrated KAN with Conv2dLSTM, mirroring a shallow network constructed using NCP neurons to facilitate performance comparison [12]. Sparsity is a defining feature of NCP neurons [11]. Accordingly, a learnable sparsity mechanism was incorporated into the KAN architecture to evaluate its impact on model performance. Integrating learnable sparsity into the KAN network enables the the evaluation of its impact on KAN neurons, aiming to enhance both model performance and efficiency.

**Table 1:** Comparsion between different networks with MLP as the baseline (*** : Superior; **: Moderate; *: Inferior)

| Network's characteristic | MLP | NCP<sup>[15, 11]</sup> | KAN<sup>[14]</sup> |
| --- | --- | --- | --- |
| Learnable activation function | * | * | *** |
| High performance | * | *** | *** |
| Reduced parameters | * | *** | *** |
| Interpretability | * | ** | *** |
| Efficiency | * | *** | *** |
| Spline based | * | * | *** |

**Figure 1.**
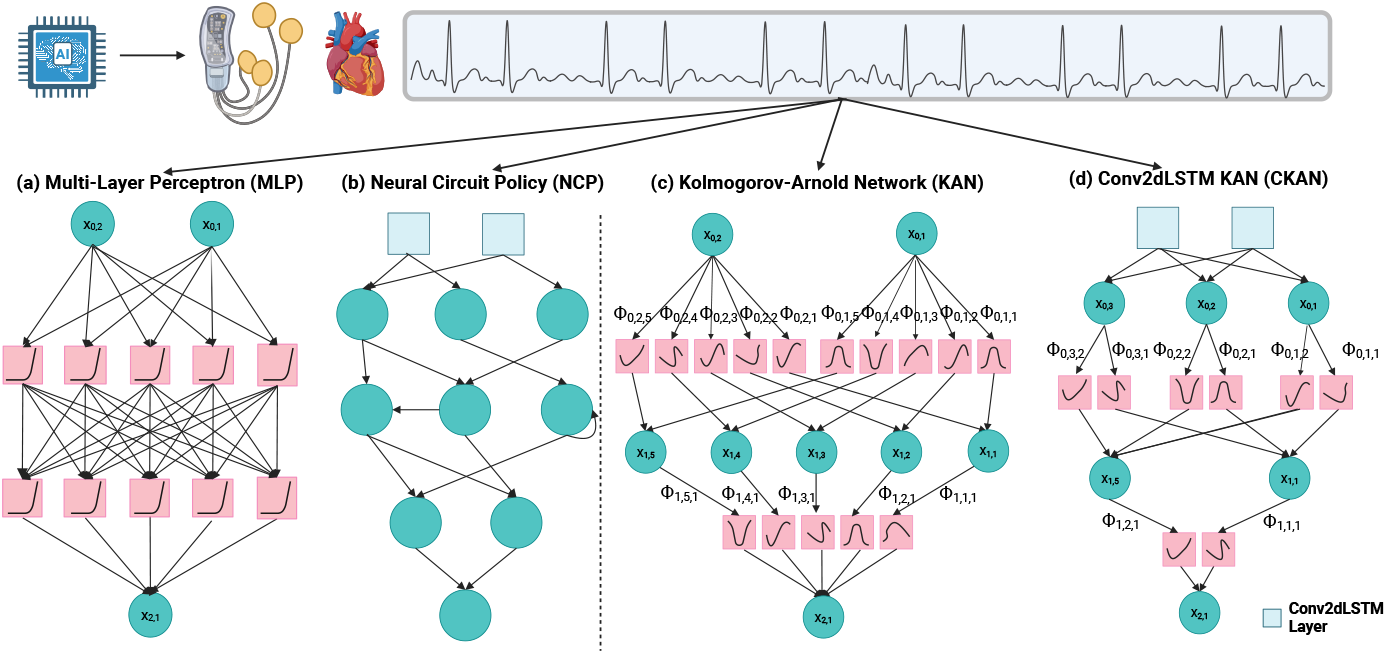
The figure illustrates four network architectures built from three types of robust neurons, highlighting their potential applications in TinyML on resource-constrained devices. (a) A traditional Multilayer Perceptron (MLP) with fixed activation functions, which is widely used in models. (b) A shallow Neural Circuit Policy (NCP) based network that emulates biological neural circuits, capable of TinyML applications. (c) A KAN network capable of learning activation functions, and (d) a shallow KAN-based network mirroring (b) with a single Conv2dLSTM layer. (a) Depicts a standard MLP with predetermined activations. (b) An NCP-based model that emphasizes modularity, information flow, and task-specific adaptability. NCP neurons are powerful units with demonstrated potential for TinyML applications. (c) Shows a KAN network that learns and adjusts activation functions based on input data, enhancing flexibility and performance. (d) Illustrates a shallow network mirroring the NCP based model design, incorporating a Conv2dLSTM layer connected to a small set of KAN neurons. Models (c) and (d) are the proposed networks in this study, evaluated for their potential applicability in TinyML.

### 1.1 Background

In the field of abnormality detection, several notable models have been developed. Petmezas et al. [16] introduced a Hybrid Convolutional Long Short-Term Memory Neural Networks (CNN-LSTM) Network with a sensitivity of 97.87% and a specificity of 99.29% for ECG heartbeat classification. A model by Gupta et al. [17] combined fractional wavelet transform, Yule-Walker autoregressive analysis, and Principal Component Analysis (PCA), achieving a mean square error of less than 0.2%, a detection accuracy of 99.89%, and an output Signal-to-Noise Ratio (SNR) of 25.25 dB. The model proposed by Chen et al. [18] utilized five Convolutional Neural Network (CNN) blocks, a bidirectional Recurrent Neural Network (RNN) layer, an attention layer, and a dense layer, achieving an F1-score of 0.84 in ECG detection. Huang et al. [19] introduced a fast compression residual CNN, achieving an average accuracy of 98.79% for ECG classification. Additionally, a shallow Diagonal State Space Sequence (S4D) model developed by Huang et al. [20] for ECG achieved a robust F1-score of 0.81 and demonstrated high robustness with input data. CNN-based AI models can also achieve expert-level accuracy in differentiating wide QRS complex tachycardias [21, 22].

Recent studies have also applied NCP models to ECG signal processing. For example, Huang et al. [12] presented two models, Conv2dLSTM-Liquid Time-Constant network (CLTC) and Conv2dLSTM-Closed-Form Continuous-time neural network (CCfC). Both models were evaluated on the ECG datasets, showing impressive generalization and robustness capabilities [12]. KANs have emerged as a novel approach in neural network architecture, inspired by the Kolmogorov-Arnold representation theorem. Unlike traditional MLPs using fixed node functions, KANs utilize learnable activation functions on network edges, enhancing flexibility and interpretability [14]. The potential of KANs in ECG analysis is underscored by their ability to handle complex signal patterns, which is essential for accurate cardiac abnormality detection.

While MLPs and their variants have been widely used for ECG signal analysis, introducing KANs represents a significant advancement. By leveraging the unique properties of the Kolmogorov-Arnold representation theorem, KANs offer a promising alternative that could enhance the accuracy, interpretability, and efficiency of ECG analysis models, ultimately improving cardiac outcomes.

## 2 Prerequisite

### 2.1 Kolmogorov-Arnold Networks (KANs)

KANs represent a significant advancement over traditional MLPs by utilizing learnable activation functions on edges rather than fixed activation functions on nodes [14]. This approach potentially allows KANs to achieve higher accuracy and interpretability, making them suitable for various applications, including time series forecasting and ECG signal classification.

### 2.2 Characteristics of KANs

KANs are based on the Kolmogorov-Arnold representation theorem, which states that any multivariate continuous function can be represented as a finite composition of continuous univariate functions and addition operations [14]. This theorem forms the foundation of KANs, enabling them to replace traditional linear weights with spline-parametrized univariate functions. The key characteristics of KANs include:

- **Learnable Activation Functions:** KANs place learnable univariate activation functions on the network’s edges, enhancing the model’s flexibility and accuracy.
- **Spline-based Representation:** Using spline functions allows KANs to adapt to the data dynamically, providing more accurate representations.
- **Improved Efficiency and Interpretability:** KANs achieve superior performance with fewer parameters than MLPs, and the learnable functions can be visualized for better interpretability.

The equation 1 represents the fundamental operation of a KAN layer.

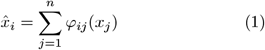

where 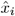 is the activation value at node *i* and *φ*_*ij*_ is the learnable activation function on the edge from node *j* to node *i*.

KAN introduces a flexible, learnable activation function *ϕ*(*x*) that combines both neural and spline-based components, allowing the model to adapt its nonlinearity based on data.

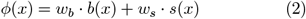

Equation 2 defines the activation *ϕ*(*x*) as a weighted combination of a basis function *b*(*x*), and *s*(*x*), a data-adaptive spline function. The weights *w*_*b*_ and *w*_*s*_ are learnable parameters that control the contribution of each component.

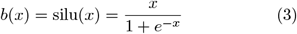

Equation 3 defines *b*(*x*) as the SiLU (Sigmoid Linear Unit) activation function, which provides smooth, differentiable nonlinearity and is known for its effective performance in deep learning due to its non-monotonic shape.

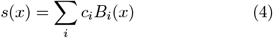

Equation 4 defines *s*(*x*) as a spline-based function, where *B*_*i*_(*x*) are spline basis functions (e.g., B-splines), and *c*_*i*_ are learnable coefficients. This term introduces localized, interpretable, and highly flexible adjustments to the activation behavior.

Together, these equations enable KAN to learn activation functions from data, blending global smoothness from SiLU with local adaptability from splines.

### 2.3 Mathematical Formulation of KANs

KANs leverage the Kolmogorov-Arnold representation theorem to decompose a high-dimensional function into a sum of univariate functions. This decomposition can be expressed as Equation 5

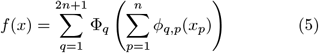

Here, φ_*q*_ and *ϕ*_*q,p*_ are univariate functions parameterized as splines, and *x*_*p*_ represents the input features. This formulation allows KANs to effectively capture both the compositional structure and univariate functions, providing a powerful framework for function approximation.

### 2.4 Application of KANs to ECG Signal Detection

In this paper, we demonstrate the significance of KANs for efficient and effective bio-signal processing by applying them to ECG abnormality classification, using two independent out-of-distribution 12-lead ECG datasets (see Sec. 3). These two datasets, one with eight classes and another with six, have four overlapping classes on which we performed our out-of-sample tests. KANs can achieve higher accuracy in detecting cardiac rhythm abnormalities due to their ability to dynamically learn and adapt activation functions. This flexibility allows KANs to model ECG signals more effectively. This efficient architecture outperforms traditional MLPs with fewer parameters, ideal for resource-limited settings. The learnable spline-based activation functions provide a more interpretable model. Clinicians can visualize these functions to understand how the model processes ECG signals. KANs are particularly adept at capturing non-linear relationships in ECG data, which are common in physiological signals. This capability leads to more robust and accurate classification.

## 3 Datasets

The datasets used in this study are identical to those employed in the previous work by Huang et al. [12], enabling a comprehensive comparison between the results of this study and prior literature in the context of TinyML applications, where models are expected to operate on resource-constrained devices. While Huang et al. [12] provides a detailed breakdown of the datasets, Table 2 presents a concise summary of their key parameters.

**Table 2:** Comparison of the TNMG and CPSC Datasets.

| Name | TNMG | CPSC |
| --- | --- | --- |
| Source | Brazil, large telehealth network | China, challenge dataset |
| Full Dataset Size | 2,322,513 annotated 12-lead ECGs | 6,877 12-lead ECGs (after filtering) |
| Sampling Rate | 400 Hz (resampled to 500 Hz for compatibility) | 500 Hz |
| Length Standardization | 4,096 readings per ECG | Standardized to 4,096 readings per ECG |
| No. of Abnormalities | 6 (AF, LBBB, 1dAVb, RBBB, ST, SB) | 8 total, 4 overlapping (AF, LBBB, 1dAVb, RBBB) |
| Subset Used in this Study | Balanced subset: 21,000 samples (3,000 per abnormality + 3,000 normal) | All 6,877 ECG recordings |
| Annotations | Clinician-verified | Expert-labeled |
| Population Characteristics | Broad population coverage, higher proportion of older patients, balanced gender and age distribution | Higher male proportion, older age distribution dominant, slight imbalance in abnormalities (fewer LBBB cases) |
| Main Usage in this Study | Training and validation | Independent test set (out-of-distribution evaluation) |

The Telehealth Network of Minas Gerais (TNMG) dataset includes six types of cardiac abnormalities: Atrial Fibrillation (AF), Left Bundle Branch Block (LBBB), First-Degree Atrioventricular Block (1dAVb), Right Bundle Branch Block (RBBB), Sinus Tachycardia (ST), and Sinus Bradycardia (SB) [4]. In contrast, the Chinese Physiological Signal Challenge 2018 (CPSC) dataset comprises eight abnormalities: AF, LBBB, 1dAVb, RBBB, Premature Atrial Contraction (PAC), Premature Ventricular Contraction (PVC), ST-segment Elevation (STE), and ST-segment Depression (STD) [23].

## 4 Methods

Our primary goal is to assess the effectiveness of KANs in processing ECG data, aiming to develop a hardware-friendly solution. To achieve this, we engineered a compact architecture that markedly reduces computational demands while upholding accuracy. Following the architecture’s development, we fed data into the model for training and validation. Then, we assessed its performance in terms of generalizability to new, unseen data and its robustness. Furthermore, we explored the network’s capabilities in handling single-lead ECG data, inching closer to potential wearable applications.

### 4.1 Data Preprocessing

Several preprocessing steps are necessary to ensure signal quality and consistency before applying the Short-Time Fourier Transform (STFT) to ECG data. These steps include bandpass filtering, wavelet denoising, and normalization.

By using bandpass filtering between 0.5 Hz and 40 Hz, removes noise outside the desired frequency range, enhancing signal quality by eliminating low-frequency drift and high-frequency noise. Wavelet denoising further cleans the ECG signals by decomposing them into different frequency components and reconstructing them while removing noise. The wavelet used is Daubechies wavelet, which is suitable for analysing signals with localised features [24]. Finally, min-max normalization scales the ECG data to a specified range, ensuring equal contribution of all features and preventing any single feature from dominating the analysis. Min-max normalization is believed to enhance the model’s generalizability [25].

The STFT is a powerful tool for analyzing non-stationary signals like ECG data, providing time-frequency representations that enable the detection of transient features that might be missed by traditional Fourier Transform methods [26, 27, 28]. By dividing the signal into overlapping segments and applying the Fourier Transform to each segment, STFT creates a two-dimensional representation with time and frequency dimensions. The window size and overlap choice are crucial for resolution and accuracy [29].

STFT has been used effectively in various studies to enhance ECG abnormality detection and classification. For example, Acharya et al. [30] used STFT to detect arrhythmias with improved performance compared to traditional methods. Similarly, De Chazal and Reilly [31] used STFT for automated ECG beat classification, achieving high accuracy in distinguishing between normal and abnormal heartbeats.

### 4.2 Model Architectures

With our focus on understanding the effectiveness of KANs, we aimed to keep the network size small. In this study, we designed and evaluated four distinct model architectures to assess their efficacy in detecting abnormalities in ECG signals. These architectures were designed with varying levels of complexity to investigate the performance trade-offs linked to different network depths and sizes. The models included:

- A single hidden layer with 64 neurons.
- A two hidden layers with 32 neurons per layer.
- A four hidden layer with 16 neurons per layer.
- A single hidden layer with 128 neurons.

Our primary aim was to utilize a network featuring an intermediate architecture of 64 neurons. We will then identify the best-performing networks within this architecture.

As illustrated in Fig. 2a), all the proposed architectures are built upon the foundational principles of KANs, employing learnable activation functions on network edges rather than fixed node functions. This design choice amplifies flexibility, adaptability, and interpretability, qualities crucial for medical applications where comprehending model decisions is paramount. The training process for these models entailed using ECG data for cardiac abnormality detection. These models underwent training to maximize performance and minimize losses. Their performance will be assessed using various metrics to understand the model’s overall performance comprehensively. The KAN model consists of a single hidden layer with 64 neurons, each using a learnable activation formed by combining a Sigmoid Linear Unit (SiLU) base function and B-spline transformations over adaptive grids. This setup enables expressive, data-driven activations. The model was trained for 50 epochs using the Adam optimizer, with regularization via an *ℓ*_1_-like penalty and an entropy term to improve sparsity and generalization. To handle overlapping pathologies, the problem was framed as a multi-label classification task. The model employed six sigmoid-activated output neurons, each representing a distinct label, with a threshold of 0.5 applied to determine binary predictions. The Binary CrossEntropy with Logits loss was used, which is well-suited for multi-label settings.

**Figure 2.**
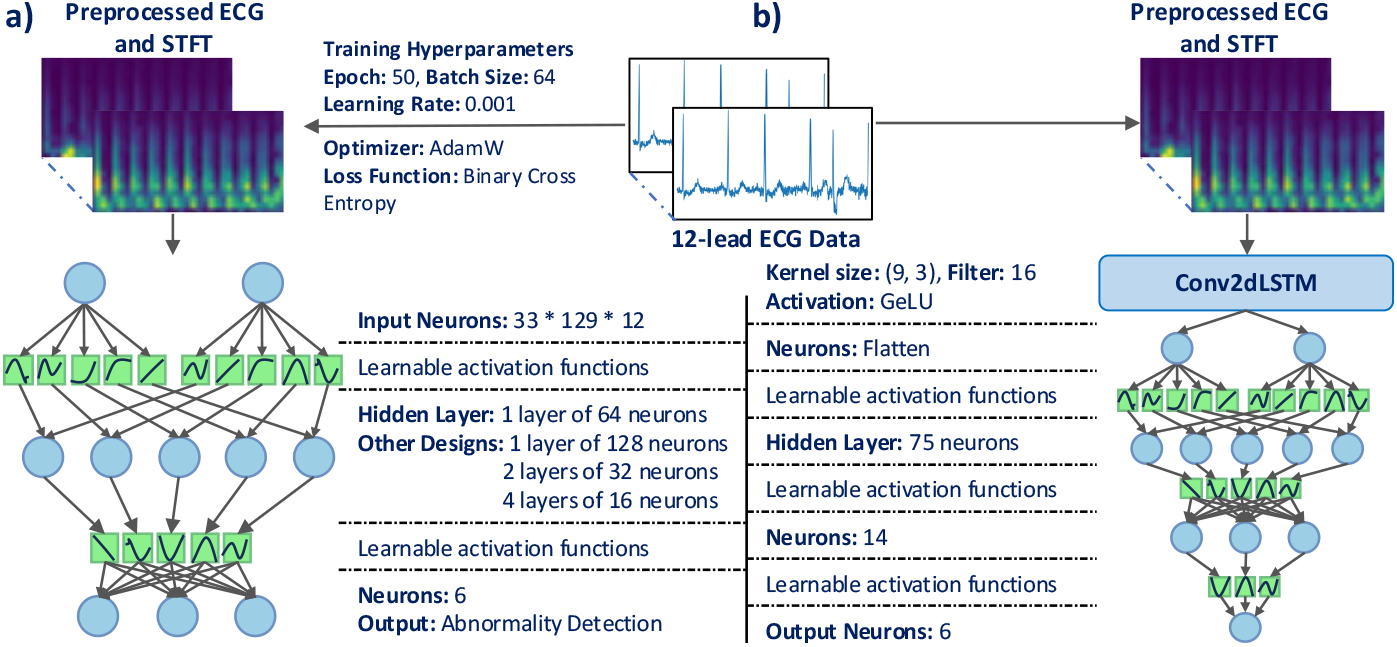
The figure illustrates the two models developed in this work. (a) The KAN model architecture consists of KAN neurons tailored for STFT-processed ECG data. The input layer comprises 33 × 129 × 12 neurons, matching the STFT pre-processed ECG data points. It then passes through intermediate layers with 64 neurons, with additional intermediate architectures also tested. Finally, six output neurons are corresponding to the six abnormalities being detected. (b) The CKAN model, a shallow network comprising KAN neurons and a single Conv2dLSTM layer, closely follows the design proposed by Huang et al. [12], which employed NCP neurons.

As part of our exploration of KAN neurons, we also developed a shallow and lightweight model based on KAN neurons (CKAN) illustrated in Fig. 2b). Inspired by the work of Huang et al. [12], which demonstrated that a simple network combining NCP neurons with a single Conv2dLSTM layer can deliver strong performance in TinyML settings, we adopted their architecture as a baseline. Using this foundation, we constructed a comparable shallow network with KAN neurons to assess its performance and potential for future TinyML applications on resource-constrained devices.

### 4.3 Evaluation Metrics

In evaluating model performance, key metrics such as precision, recall, and their combined measure, the F1score, play pivotal roles. Precision gauges the accuracy of positive predictions, while recall indicates the proportion of actual positives correctly identified, offering insights into the model’s performance trade-offs.

When dealing with binary classification, metrics like the AUROC (Area Under the Receiver Operating Characteristic curve) measure a model’s efficacy in determining negative and positive cases through various threshold levels. Conversely, the AUPRC (Area Under the Precision-Recall Curve) emphasizes the model’s accuracy in pinpointing positive cases, which is particularly useful in handling imbalanced datasets.

Metrics such as F1-score, precision, recall, AUROC, and AUPRC are crucial for evaluating machine learning model performance, especially in tasks like abnormality detection.

## 5 Experiment

### 5.1 The Training Process

In the experimental phase of our study, we utilized the TNMG subset data as the primary training dataset for our KAN models. This dataset provided diverse ECG recordings encompassing various cardiac abnormalities. The models underwent training for 50 epochs with a batch size of 64. We utilized the AdamW optimizer with a weight decay of 10^*−*4^ and employed binary crossentropy loss. A scheduler (ExponentialLR) was also implemented with a gamma of 0.9.

While the dataset seems balanced overall, there remains an imbalance between total positive and negative cases due to the multi-labeled nature of abnormalities. This means a person can have more than one abnormality. To rectify this skewness and enhance the model’s ability to detect positive cases, a technique called repetition- of-4 was implemented. This approach involves replicating all data instances with positive cases four times within the training data. This technique has improved the model’s performance in detecting positive cases [12]. While Huang et al. [12] has used repetition-of-2 in their work, repetition-of-4 was tested in this case providing better performance.

### 5.2 In-sample Training and Validation Procedures

For assessing model performance, we allocated a dedicated dataset comprising 20% of the TNMG subset to serve as validation data. These data points were deliberately excluded from the training phase to prevent information leakage and to provide an unbiased benchmark for evaluation. By withholding this portion of the dataset, we ensured that the validation process reflected the model’s ability to generalize to unseen data rather than simply memorizing patterns from the training set. This separate validation strategy allowed for a fair and consistent comparison of the individual models’ performances, enabling us to identify strengths and weaknesses in their predictive capabilities and to make informed decisions regarding subsequent model refinement.

### 5.3 Evaluation on Unseen Data

We evaluated our model’s performance using the CPSC dataset, which closely represents real-world scenarios. This assessment involved comparing the models’ predictions against data labels using a variety of performance metrics. This in-depth analysis will help us grasp the models’ strengths and limitations, providing valuable insights into their effectiveness. Additionally, these findings will form the basis for identifying potential areas to enhance ECG analysis. We specifically selected the four abnormalities in both CPSC and TNMG for comparison.

### 5.4 Model Robustness

The experiments will be systematically conducted by incrementally increasing the number of randomly removed channels. In the repeated experiments, a defined number of ECG channels were randomly removed from the signal, ranging from 1 to 6: the first experiment removed 1 lead, the second removed 2, and so on, up to 6 leads. This approach allows us to assess the model’s resilience to varying degrees of missing input data. These experiments aim to understand the model’s capacity to sustain accuracy across a spectrum of scenarios. We will utilize a range of performance metrics to assess its performance objectively. These metrics will facilitate a thorough evaluation and comparison of the model’s efficacy under different conditions. The findings from these assessments will help identify areas for improvement and offer direction for future enhancements.

### 5.5 Model for Single-Lead ECG

In addition to the evaluations conducted with the 12- lead ECG data, we designed an experiment to assess the model’s performance using data with only a single lead. Specifically, we utilized Lead II data to train the KAN model. We selected Lead II for the single-lead experiment because it is the most commonly used lead for ECG monitoring and gives an electrical view that closely aligns with the heart’s natural depolarization vector, yielding clear P, QRS, and T wave morphology [32]. This experiment aimed to investigate whether the model could maintain its performance when trained with data of just one lead, thereby evaluating its potential for use in scenarios where only single-lead ECG data is available. The training procedure, hyperparameters, and validation methods remained consistent with those used in the 12- lead ECG experiments to ensure a fair comparison. The only change we made was reducing the input layer from 33 × 129 × 12 to 33 × 129 × 1, reflecting the reduction in input data.

### 5.6 Model Interpretability

The KANs model is known for its interpretability, as each edge’s activation function is accessible. To illustrate this, we plot the top 3 activation functions connected to each of the 6 decision neurons (each corresponding to a classification). Importance was based on spline coefficient magnitudes: larger values indicate stronger non-linear influence, while near-zero values suggest minimal effect, highlighting which inputs captured more complex patterns.

We also assess the model by generating a saliency map, which assigns each input element an importance score based on the gradient of the output concerning the input, indicating how sensitive the prediction is to changes in that region. By visualizing these scores as a heatmap, saliency maps produce intuitive insights into where the model “looks” when making decisions. This enhances interpretability by revealing influential features, helping users debug unexpected behavior, uncover biases, and build trust in the model’s predictions.

### 5.7 KAN-Based Shallow TinyML Network

To enable a meaningful comparison and to better understand the capabilities and limitations of KAN neurons, we constructed a shallow network consisting of KAN neurons and a single Conv2dLSTM layer. The architecture closely follows the design proposed by Huang et al. [12], where a shallow network with NCP neurons and a Conv2dLSTM layer was developed for TinyML applications. While our setup mirrors this prior work, minor adjustments were introduced. Specifically, the Conv2dLSTM layer employs a kernel size of (9, 3) with 16 filters and GeLU activation. Its output is connected to 75 recipient KAN neurons, which in turn connect to 14 processor KAN neurons, and finally project to 6 output neurons corresponding to six abnormalities. Hyperparameters were selected following standard practice, including a learning rate of 0.001, Binary Cross-Entropy loss with early stopping, the AdamW optimizer, and a ReduceLROnPlateau scheduler with a patience of 5 and a reduction factor of 0.5. The model will be trained and validated on the TNMG dataset and evaluated on the CPSC dataset to assess its generalization capability. The model will additionally be trained using only Lead II ECG data from the TNMG dataset to evaluate its performance when limited to a single lead. The results obtained from this shallow model provide a basis for comparing different neuron types and justify the potential application of KAN neurons in TinyML, warranting further analysis and development. The CKAN has undergone in-sample training and validation on TNMG, out-of-sample generalization on CPSC, and single-lead ECG testing on TNMG.

### 5.8 Integrating Learnable Sparsity

One of the key strengths of NCP neurons is their sparse connectivity, which enhances efficiency and biological plausibility [11]. Sparsity also enables model pruning, reducing complexity for deployment on TinyML devices. Inspired by this, we investigate integrating sparsity into the KAN network during training to assess its effect on inference efficiency and predictive performance under hard sparsity (pruning weak connections).

To improve computational efficiency and generalization, we introduced learnable sparsity within the KAN layers of the CKAN model. A trainable mask, derived from a parameter tensor passed through a sigmoid function, modulates input–output connections, suppressing less relevant ones. The mask is optimized jointly with other parameters via AdamW, guided by a composite regularization combining L_1_, entropy, and mean-mask (M) penalties, weighted by a hyperparameter *λ* as presented in Equation 6. During inference, hard thresholding (mask less than 0.5 is set to 0) enforces a fixed sparse structure. This allows the model to adaptively retain only the most relevant connections, balancing performance and efficiency.

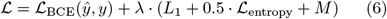

The CKAN model with learnable sparsity was trained on the TNMG dataset using the same hyperparameters and then evaluated on the CPSC dataset for generalization. All hyperparameters were kept constant, except for the sparsity regularization coefficient *λ*, which was set to 1 × 10^*−*3^ and 1 × 10^*−*2^ to study its impact. A larger *λ* encourages stronger sparsity in the network.

## 6 Results

In this section, we will comprehensively analyze the models’ performance, specifically focusing on utilizing KAN models for training and evaluation. Our discussion will explore the models’ generalizability capabilities and robustness. Additionally, we will showcase the model’s potential for single-lead ECG data analysis.

### 6.1 Training with Different Architecture

As we seek to understand the effectiveness of KAN neurons and explore their potential for use in hardware applications that demand compact network sizes and fewer parameters, we are conducting a study focused on small networks. Accordingly, we have designed a network comprising 64 neurons evenly distributed across 1, 2, and 4 hidden layers. This design encompasses a model with a single intermediate layer of 64 neurons, two intermediate layers of 32 neurons each, and four intermediate layers of 16. We trained these models using 50 epochs, and Fig. 3 illustrates the training process of these models.

**Figure 3.**
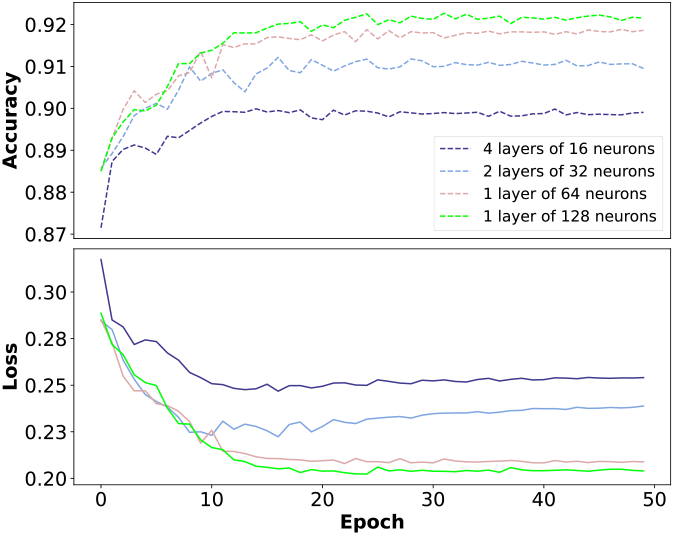
The training processes of various KAN architectures show increasing accuracy and decreasing loss throughout. However, discrepancies can be observed among different network architectures.

We can observe that with the same number of neurons, the model trains more effectively with a single layer of 64 neurons, as evidenced by the lower stabilization of loss and higher accuracy. For completeness, we also tested a model with an intermediate layer of 128 neurons, and it’s noticeable that as the number of neurons increases, the model’s performance improves.

While a more complex network yields better performance, we opted for the model with one intermediate layer of 64 neurons for further analysis. Our focus is on understanding the effectiveness of these neurons in ECG applications, with an eye toward future hardware implementations.

### 6.2 In-Sample Performance Evaluation

This research study focuses on training the proposed KAN model using the TNMG subset dataset, as detailed earlier in this paper. The TNMG subset dataset serves as the primary resource for developing and evaluating the models in this investigation. The model’s validation results on in-sample pre-processed data are summarized in the top-left of Table 3. The model exhibits robust performance, achieving an F1-score of 0.75. Moreover, the AUROC value stands at 0.95, with an AUPRC value of 0.84.

**Table 3:** Validation and Generalization Results (R: Recall, P: Precision)

| KAN |  |  |  |  |  | CKAN |  |  |  |  |
| --- | --- | --- | --- | --- | --- | --- | --- | --- | --- | --- |
| Class | R | P | F1 | AUROC | AUPRC | R | P | F1 | AUROC | AUPRC |
| **In-Sample Validation on TNMG dataset** |  |  |  |  |  |  |  |  |  |  |
| 1dAVb | 41.3% | 52.6% | 46.3% | 80.6% | 46.6% | 60.5% | 63.4% | 61.9% | 89.2% | 67.1% |
| RBBB | 77.8% | 83.9% | 80.7% | 95.1% | 88.8% | 88.3% | 93.0% | 90.6% | 98.2% | 95.1% |
| LBBB | 87.3% | 91.2% | 89.2% | 98.8% | 95.7% | 92.9% | 92.7% | 92.8% | 99.2% | 97.0% |
| SB | 87.7% | 88.6% | 88.2% | 98.7% | 94.4% | 88.9% | 87.4% | 88.1% | 98.9% | 94.1% |
| AF | 41.6% | 54.1% | 47.1% | 81.8% | 50.8% | 69.7% | 71.7% | 70.7% | 93.3% | 76.3% |
| ST | 86.1% | 88.3% | 87.2% | 98.3% | 93.4% | 93.8% | 90.6% | 92.1% | 99.4% | 97.4% |
| Average | 71.2% | 78.8% | 74.8% | 94.6% | 84.1% | 83.0% | 84.1% | 83.5% | 97.4% | 90.9% |
| **Out-of-Distribution Generalization** |  |  |  |  |  |  |  |  |  |  |
| 1dAVb | 41.7% | 34.0% | 37.5% | 78.1% | 34.0% | 79.1% | 50.5% | 61.6% | 91.5% | 71.8% |
| RBBB | 54.1% | 85.2% | 66.2% | 80.3% | 74.5% | 64.9% | 91.0% | 75.8% | 87.0% | 82.3% |
| LBBB | 78.7% | 86.0% | 82.2% | 97.3% | 86.2% | 83.8% | 82.4% | 83.1% | 97.0% | 87.3% |
| AF | 59.3% | 42.4% | 49.5% | 78.9% | 48.3% | 83.2% | 58.0% | 68.4% | 92.6% | 75.5% |
| Average | 67.3% | 67.4% | 62.2% | 83.6% | 60.7% | 77.8% | 70.5% | 72.2% | 92.0% | 79.2% |
| **In-sample Validation on Single-lead ECG Data** |  |  |  |  |  |  |  |  |  |  |
| 1dAVb | 41.0% | 58.1% | 48.0% | 86.1% | 55.1% | 55.1% | 69.5% | 61.5% | 89.1% | 65.7% |
| RBBB | 61.4% | 69.2% | 65.1% | 87.7% | 70.8% | 71.7% | 78.0% | 74.7% | 92.7% | 81.6% |
| LBBB | 67.7% | 79.1% | 72.9% | 94.8% | 83.1% | 81.6% | 82.7% | 82.1% | 96.9% | 89.9% |
| SB | 88.0% | 86.2% | 87.1% | 98.5% | 93.2% | 89.7% | 84.2% | 86.9% | 98.6% | 92.4% |
| AF | 36.3% | 50.9% | 42.4% | 81.4% | 46.3% | 57.1% | 73.6% | 64.3% | 91.9% | 72.8% |
| ST | 85.7% | 89.3% | 87.5% | 98.1% | 93.4% | 91.5% | 88.5% | 89.9% | 99.1% | 95.5% |
| Average | 63.6% | 73.9% | 68.3% | 92.7% | 77.8% | 74.6% | 80.2% | 77.3% | 95.5% | 85.1% |

These results highlight that even a single hidden layer KAN network model performs robustly, indicating significant potential for ECG analysis tasks. The observed performance metrics underscore the importance of data pre-processing methods tailored to the specific application.

From the top-left of Table 3, it can be seen that the performance of the model on detecting AF and 1dAVb is generally harder in terms of F1-scores. This could be due to the following reasons:

- Irregularity of AF: AF causes an irregular heart rhythm with no distinct P waves, making it more challenging to identify. The irregular intervals between beats make it harder to develop consistent patterns for machine learning models.
- Subtlety of 1dAVb: 1dAVb is often asymptomatic and presents as a prolonged PR interval on an ECG. This lengthening can be subtle and is easily missed without precise measurements.

To improve the performance, the F1-score can be adjusted by using different threshold values in determining positive and negative cases. Also, the simple KANs model could still be improved in later development as this study is only focused on testing the capability of the simplest KANs neurons.

The proposed CKAN model, consisting of a single Conv2dLSTM layer combined with KAN neurons and closely mirroring the architecture introduced by Huang et al. [12] for TinyML applications, was evaluated as follows. After being trained on the TNMG dataset, the model was assessed on the in-sample validation set, with the results summarized in the top-right of Table 3. The results show relatively strong performance, with high F1 and AUROC scores, suggesting that the model may be effective.

### 6.3 Model Generalization

Our evaluation strategy involved applying our trained models from the TNMG subset to make predictions on the CPSC dataset, as outlined in the data methodology. This assessment was designed to ascertain how effectively our models could handle new data instances from the CPSC dataset, which may possess inherent differences compared to the training dataset. Notably, the CPSC dataset encompasses eight distinct types of abnormalities, with only four overlapping with those in the TNMG dataset.

Analyzing the KAN model’s performance on the CPSC dataset provides valuable insights into its ability to transfer learned knowledge to new data domains. This examination is a critical benchmark for assessing the model’s generalization capabilities beyond its original training data, demonstrating its adaptability to a broader range of clinical scenarios.

Middle-left of Table 3 outlines the model’s generalization performance, achieving an F1-score of 0.62, an AUROC of 0.84, and an AUPRC of 0.61. While a decrease in performance is noticeable, the drop remains within an acceptable range based on our experience. Considering the simplicity of the architecture we’ve introduced, there is significant potential for further improvement with advancements in research in this novel network. These results highlight the potential of the KAN network model in ECG analysis, indicating its viability for real-world applications. Given that this network comprises solely KAN neurons, there is considerable room for enhancing the performance of this model with future research into the network.

To evaluate the generalization ability of the CKAN model on unseen data, we performed inference on the CPSC dataset, and the results are summarized in the middle-right of Table 3. While the performance declined, which is not uncommon in generalization tests, the drop remained within an acceptable range. Moreover, performance could likely be recovered with minimal fine-tuning on the new dataset. The CKAN model demonstrates relatively fast training, requiring only about 30 to 60 minutes on an NVIDIA V100 Graphics Processing Unit (GPU). This efficiency supports its suitability for finetuning and continual learning tasks. With a compact size of just 6.5 MB, the model is highly lightweight and well-suited for TinyML applications.

### 6.4 Model Robustness

To evaluate the model’s efficacy, we broadened our analysis to incorporate the CPSC dataset, deliberately introducing systematic variability. Furthermore, we performed supplementary evaluations by randomly omitting a defined number of channels from the 12-lead ECG data. This deliberate alteration of input data enabled us to assess the model’s robustness and its aptitude to manage incomplete or absent input information proficiently. These experimental maneuvers significantly enhance our comprehension of the model’s adaptability and resilience in practical scenarios where data integrity might be compromised.

In Fig. 4, it can be observed that as the number of emptied leads increases, the performance metric of the models declines. While min-max normalization can improve robustness to missing channels [25], excessive missing channels can still negatively impact model performance. AUROC is relatively unaffected, indicating that the model can still discriminate between classes overall, even when conditions change. In contrast, AUPRC is more sensitive, suggesting that the model’s precision in identifying positive cases is more strongly impacted under these conditions.

**Figure 4.**
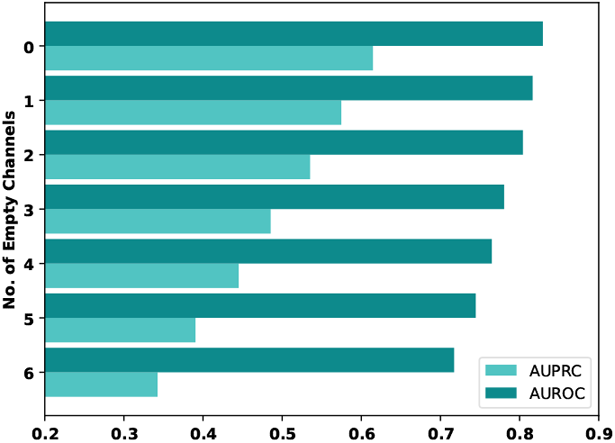
Assessing KAN model robustness using AU-ROC and AUPRC metrics across different levels of missing channels in CPSC ECG data.

### 6.5 Exploring Model Performance with Single-Lead ECG Data

As part of our investigation into the model’s capabilities, we conducted an experiment utilizing the TNMG dataset. Interestingly, for this specific experiment, we focused solely on Lead II data to train the KAN model. The detailed outcomes of this experiment can be found in the bottom-left of Table 3.

Contrasting the outcomes outlined in the top-left of Table 3, originating from a model trained on comprehensive 12-lead ECG data, with the results of a model exclusively trained on Lead II data reveals intriguing observations. While there’s a slight dip in the overall performance of the latter model across various abnormalities, it’s remarkable that such proficiency was achieved using data from only one lead. Despite this limitation, the model’s performance remains robust.

The CKAN model was additionally trained using only Lead II data from the TNMG dataset, and its performance was evaluated on the TNMG validation set. Bottom-right of Table 3 provides a detailed breakdown of the results. Despite using only a single ECG lead, the mirrored model performs reasonably well, demonstrating the potential of KAN neurons for TinyML applications.

### 6.6 Exploring Model Interpretability

To illustrate the key feature of the KAN model, where edges connect neurons with learnable activation func-tions, we have plotted the three most important activation functions associated with the output neurons used to detect the abnormalities, as shown in Fig. 5. This learnable activation function feature enhances the interpretability of the model by capturing non-linear relationships through adaptive activation functions.

**Figure 5.**
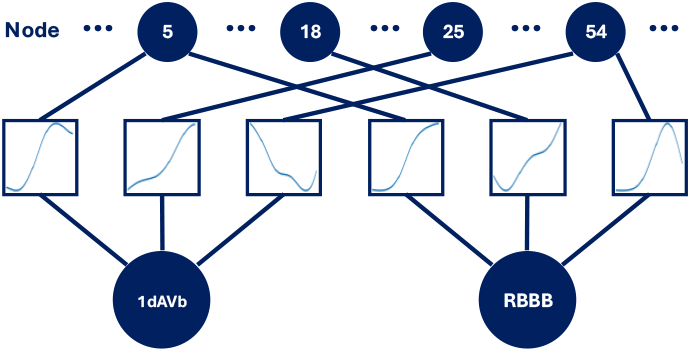
The top three most important activation functions on the edges connecting to the decision nodes (neurons) of the KAN model were identified based on the magnitudes of their spline coefficients. Larger coefficients indicate stronger nonlinear influence, while nearzero values suggest minimal contribution. The visualization highlights the most influential activation functions of the nodes linked to 1dAVb and RBBB classes.

As a demonstration, we generate saliency maps to reveal which time–frequency segments of the STFT-processed 12-lead ECGs with RBBB most drive the model’s output, spotlighting the regions exerting the greatest influence on its predictions. Figure 6 highlights which areas are most important across each lead, providing insight into the model’s focus during inference.

**Figure 6.**
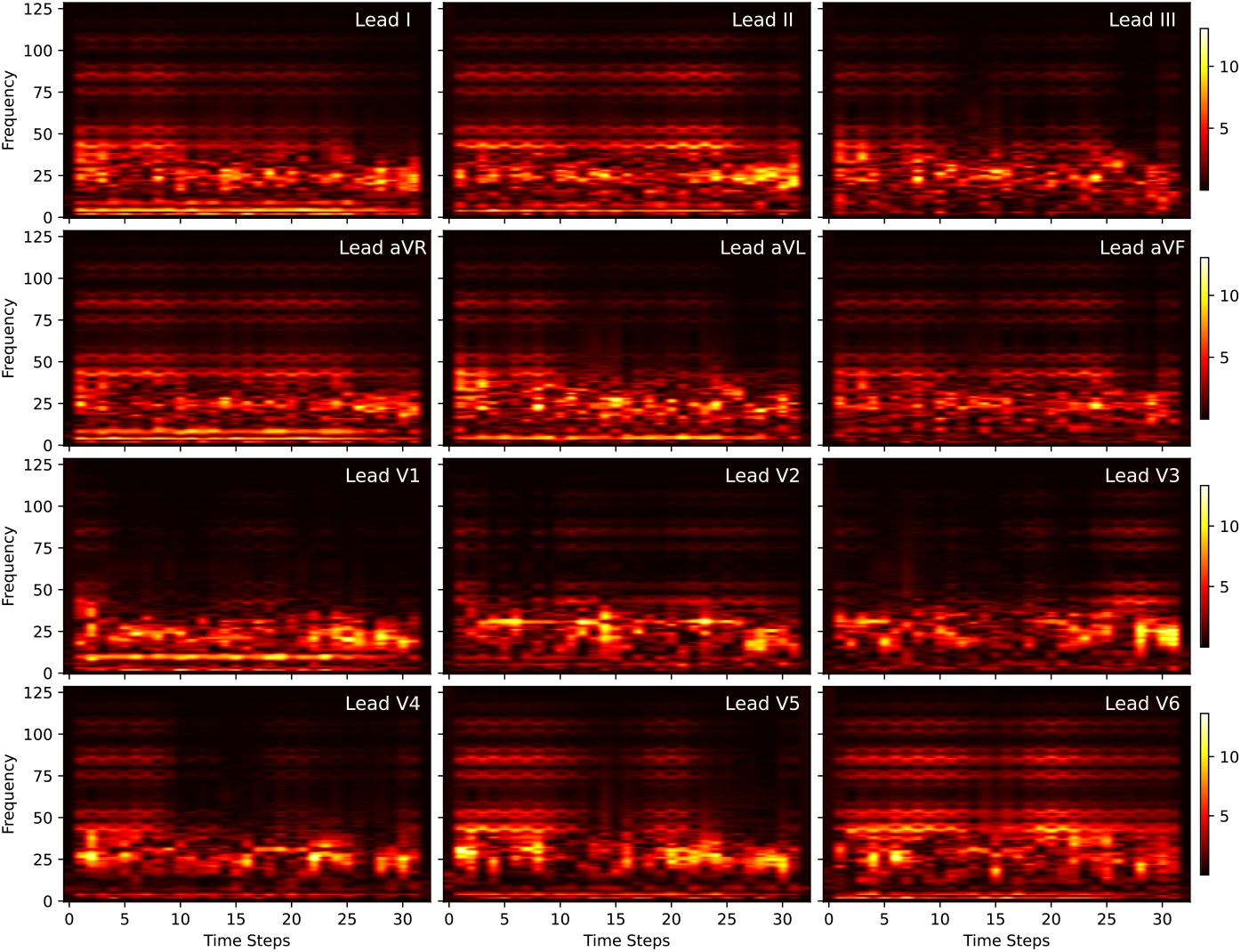
Saliency maps over STFT-transformed 12-lead ECGs with RBBB, highlighting the time-frequency regions that contributed to the KAN model’s prediction. The model consistently focused on features below 50 Hz across all 12 leads when identifying RBBB, while certain features above 50 Hz also played a notable role in the decisionmaking process. The x-axis represents the time steps across the 10-second ECG data segment.

### 6.7 Learnable Sparsity

The CKAN model with learnable sparsity was trained on the TNMG dataset. Both the soft and hard versions of the model were validated on TNMG and then evaluated on the CPSC dataset for generalization. The soft model retains the learned sparsity as is, whereas the hard model prunes connections with sparsity values below 0.5 by setting them to zero, making it more suitable for inference for TinyML applications. Two sparsity regularization coefficients, *λ* = 1 × 10^*−*3^ and *λ* = 1 × 10^*−*2^, were tested. The results for *λ* = 10^*−*3^ are presented in Table 4, and those for *λ* = 10^*−*2^ are shown in Table 5.

**Table 4:** Validation and Generalization of Sparse CKAN with *λ* = 1 *×* 10^*−*3^ (R: Recall, P: Precision)

|  | Soft |  |  |  |  | Hard |  |  |  |  |
| --- | --- | --- | --- | --- | --- | --- | --- | --- | --- | --- |
| Class | R | P | F1 | AUROC | AUPRC | R | P | F1 | AUROC | AUPRC |
| In-Sample Validation on TNMG dataset |  |  |  |  |  |  |  |  |  |  |
| 1dAVb | 68.3% | 72.0% | 70.1% | 93.3% | 75.5% | 63.4% | 69.5% | 66.3% | 91.6% | 72.4% |
| RBBB | 87.7% | 93.3% | 90.4% | 98.6% | 95.7% | 87.3% | 93.9% | 90.5% | 98.7% | 95.8% |
| LBBB | 93.0% | 92.9% | 93.0% | 99.2% | 97.5% | 91.9% | 92.7% | 92.3% | 99.1% | 97.2% |
| SB | 91.4% | 85.4% | 88.3% | 98.9% | 94.0% | 90.7% | 87.6% | 89.1% | 98.9% | 94.1% |
| AF | 72.3% | 79.1% | 75.5% | 95.8% | 83.8% | 67.9% | 79.8% | 73.4% | 95.1% | 82.5% |
| ST | 91.6% | 91.9% | 91.8% | 99.5% | 97.6% | 93.1% | 91.6% | 92.4% | 99.4% | 97.4% |
| Average | 84.5% | 86.4% | 85.5% | 98.1% | 92.8% | 82.9% | 86.8% | 84.8% | 97.9% | 92.3% |
| Out-of-Distribution Generalization on CPSC dataset |  |  |  |  |  |  |  |  |  |  |
| 1dAVb | 81.7% | 56.6% | 66.9% | 93.1% | 77.8% | 81.4% | 57.1% | 67.2% | 93.2% | 78.0% |
| RBBB | 67.1% | 90.9% | 77.2% | 89.0% | 84.4% | 67.0% | 91.2% | 77.2% | 88.9% | 84.4% |
| LBBB | 85.9% | 80.8% | 83.3% | 97.2% | 87.6% | 85.9% | 80.8% | 83.3% | 97.1% | 87.5% |
| AF | 83.9% | 63.4% | 72.2% | 93.3% | 78.0% | 82.9% | 65.0% | 72.9% | 93.1% | 78.5% |
| Average | 79.7% | 72.9% | 74.9% | 93.1% | 81.9% | 79.3% | 73.5% | 75.1% | 93.1% | 82.1% |

**Table 5:**
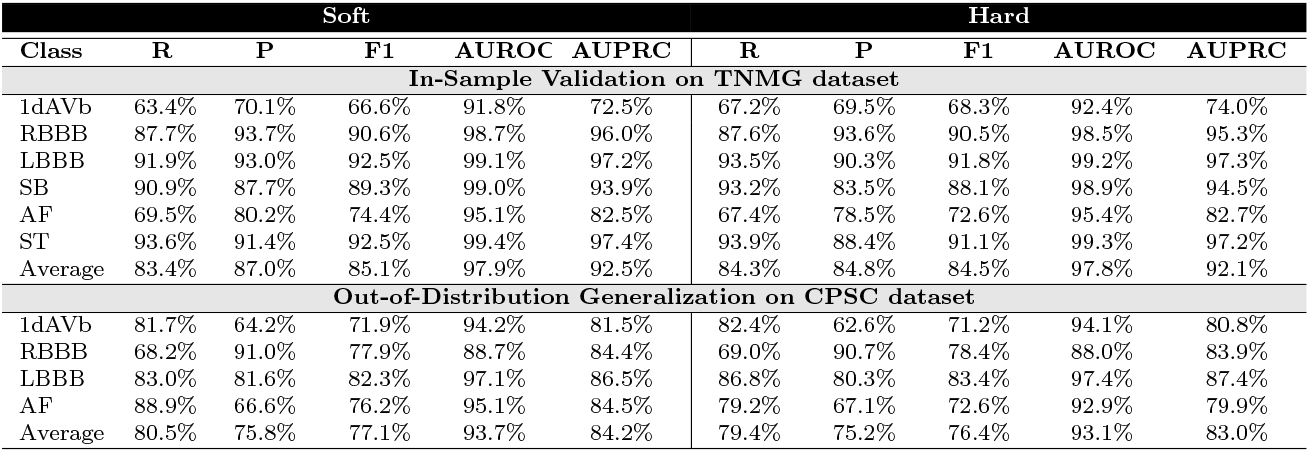
Validation and Generalization of Sparse CKAN with *λ* = 1 *×* 10^*−*2^ (R: Recall, P: Precision)

Surprisingly, the introduced learnable sparsity during training has boosted the performance of the CKAN model, particularly when *λ* is relatively large. This improvement is evident in the generalization test, where the hardened model, obtained by pruning weights with sparsity below 0.5, still maintains strong performance. These results highlight the potential of KAN for TinyML applications, especially in resource-constrained environments. The performance even surpasses that of the CKAN model trained without sparsity.

After hardening, 17.6% of the CKAN model’s weights were pruned when trained with *λ* = 1 × 10^*−*3^, and 53.5% were pruned with *λ* = 1 × 10^*−*2^, as a higher *λ* encourages greater sparsity. Despite the pruning, the KAN neurons maintained strong performance under learnable sparsity.

## 7 Discussion

This study represents the initial successful application of KAN to ECG signal analysis, particularly focusing on multi-abnormality detection. The results demonstrate promising and satisfactory outcomes. The proposed KAN models have shown strong performance and robust generalization capabilities, effectively handling situations involving incomplete or missing input data. Even a single hidden layer KAN network, with only 64 nodes in its hidden layer, exhibited good performance. This highlights the flexibility and adaptability of KAN networks, even in extremely simple structures.

Compared to traditional MLPs, KANs offer significant advantages. MLPs, while powerful, often struggle with fixed activation functions and less interpretable structures, which can hinder performance and clinical applicability [14]. In contrast, KANs leverage the Kolmogorov-Arnold representation theorem to implement learnable activation functions on the network’s edges, enhancing flexibility and interpretability. Additionally, the study compared KANs to more recent and compact models such as NCP models. NCPs, a bio-inspired neural network, have demonstrated significant success in efficient and robust ECG abnormality detection [12]. Despite these advanced capabilities, the simple single hidden layer KAN network with 64 neurons in its hidden layer performed comparably, underscoring its potential as a highly adaptable and efficient model for ECG analysis. A notable characteristic of KANs is their efficiency. The model’s compact size and quick training make it well-suited for potential deployment on hardware, especially in personalized medical devices. This efficiency supports real-time data processing and reduces power consumption, a critical factor for continuous monitoring applications.

In our evaluation, we compared CKAN and single hidden-layer KANs with NCP-based models [12], using standard performance metrics including precision, recall, F1-score, and AUROC. We implemented two variants of NCPs by utilizing two distinct neuron types: Liquid Time Constant (LTC) and Closed Form Continuous (CfC). Each was used to construct a shallow network augmented with a Conv2dLSTM layer, resulting in two models, CLTC and CCfC. A detailed comparison of their performance is provided in Table 6. We also trained an MLP model with the same architecture as the proposed KAN, but it did not yield significant results. This underscores the power and efficiency of the neurons in the KAN model. All experiments were conducted under identical conditions using the complete 12-lead ECG dataset.

**Table 6:**
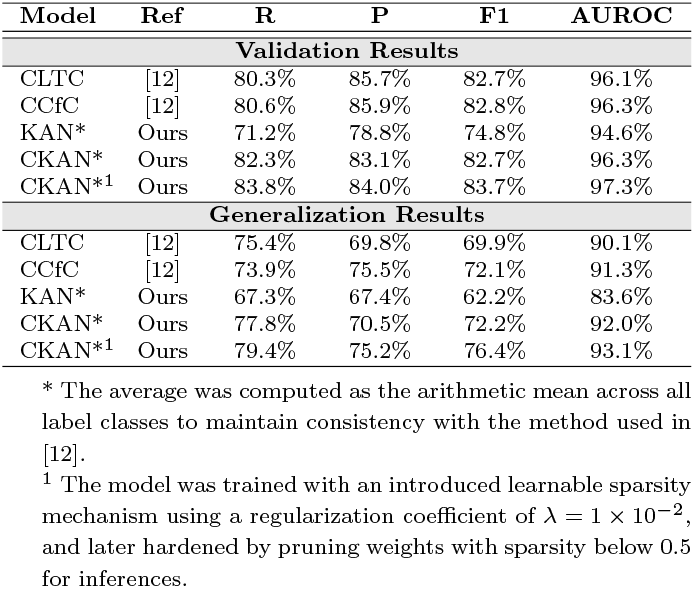
Validation and Generalization Results (R: Recall, P: Precision)

While the KAN model may lag in performance compared to other efficient neural networks like NCPs, their architecture has fundamental differences. The CCfC and CLTC models incorporate a layer of Conv2dLSTM to aid in the feature extraction process, whereas the proposed KAN model does not include additional layers for this purpose. After adding a single Conv2dLSTM layer, the CKAN model performed on par with, if not slightly better than, the CCfC and CLTC models in terms of F1 and AUROC, despite not being specifically optimized but instead simply mirrored to their structures. The mirrored structure may not represent the optimal architecture for fully leveraging the potential of KAN neurons. This suggests that KAN neurons may surpass NCP neurons in performance, and that CKAN could be a stronger candidate for TinyML applications, pending further evaluation, given that both CCfC and CLTC were originally designed as lightweight models for TinyML deployment. CKAN’s reasonable performance on single-lead ECG data further confirms its potential for TinyML applications in resource-constrained wearable devices. The training of CKAN models was also relatively efficient compared to NCP neurons, which are based on ordinary differential equations (ODEs) and generally require more computation to train [11].

After introducing a learnable sparsity scheme, a key characteristic of NCP neurons, into the KAN neurons of the CKAN model, it was surprising to observe an enhancement in model performance and generalization. Even after pruning weak sparse weights, the model still outperformed both the original CKAN model without learnable sparsity and similar architectures using NCP neurons. This result demonstrates that sparsity and KAN neurons can work synergistically, not only improving performance but also increasing efficiency for potential TinyML applications where weights are pruned for resource optimization.

### 7.1 Related Works

There is growing interest in the application of machine learning models to ECG analysis. For example, Zeleke and Bochicchio [33] introduced a model that integrates Federated Learning (FL) with KANs to enable privacy-preserving health data monitoring on the MIT-BIH arrhythmia dataset. Likewise, Zhao et al. [34] employed KAN for single-class ECG classification focused on atrial fibrillation (AF). Beyond ECG, Nimishan et al. [35] developed a KAN-based model for EEG-based emotion recognition, achieving an average accuracy of 80.79% and an F1 score of 78.63% on the GAMEEMO dataset, and an accuracy of 90.95% with an F1 score of 89.53% on the LUMED dataset. A comparison of representative works using similar datasets or targeting TinyML applications, is presented in Table 7. Unlike existing approaches, our proposed networks leverage the novel KAN framework to achieve a more streamlined architecture, demonstrating strong potential for deployment on resource-constrained edge devices.

**Table 7:** Evaluating Model Performance in Comparison to Related Work.

| Ref | Model Used | Dataset | Task | Precision | Recall | F1 | AUROC | Accuracy |
| --- | --- | --- | --- | --- | --- | --- | --- | --- |
| [36] | CNN | CPSC | Multi-label cardiac arrhythmia classification | 39.4% | 53.3% | 41.3% | 96.2% | 95.6% |
| [37] | Multi-channel multi-scale deep neural network | CPSC | Automated ECG classification | – | – | 67.34-74.73% | – | 94.1% |
| [38] | 3DECG-Net | CPSC | Multi-label cardiac arrhythmia detection | – | – | 69.4-85.7% | 81.4% | 81.6% |
| [39] | Deep representation learning | TNMG | Atrial fibrillation risk prediction from 12-lead ECG | – | – | – | 90.9% | 94.9% |
| [40] | TinyCES | MIT-BIH, PTB | ECG Classification | – | – | – | – | 97% |
| [41] | CNN | MIT-BIH | TinyML-Based Classification | – | – | – | – | 92.3% |
| Our Work | KAN | TNMG | Multi-label ECG | 78.8% | 71.2% | 74.8% | 94.6% | – |
|  |  | CPSC* | anomalies detection | 67.4% | 67.3% | 62.2% | 83.6% | – |
| Our Work | CKAN | TNMG | Multi-label ECG | 84.1% | 83.0% | 83.5% | 97.4% | – |
|  |  | CPSC* | anomalies detection | 70.5% | 77.8% | 72.2% | 92.0% | – |
| Our Work | Sparse CKAN<sup>1</sup> | TNMG | Multi-label ECG | 84.8% | 84.3% | 84.5% | 97.8% | – |
|  |  | CPSC* | anomalies detection | 75.2% | 79.4% | 76.4% | 93.1% | – |

\* The model was not trained on this dataset; instead, it was used for inference to assess its generalization ability.
<sup>1</sup> The model was trained with an introduced learnable sparsity mechanism using a regularization coefficient of $\lambda = 1 \times 10^{-2}$ , and later hardened by pruning weights with sparsity below 0.5 for inferences.

## 8 Limitations and Future Work

### 8.1 Limitations

Despite the promising results demonstrated by KANs in ECG signal analysis, several limitations need to be addressed:

- **Novelty and Integration:** KANs are a relatively new architecture and have not yet been widely tested or integrated with other advanced neural network layers such as CNNs or RNNs. This limits the current understanding of how KANs can be effectively combined with other deep-learning models to enhance performance further.
- **Code Optimization:** The existing implementations of KANs are still in the early stages and may not be fully optimized for large-scale deployment. The current codebase might lack the efficiency and robustness required for real-world applications, particularly in resource-constrained environments like wearable devices.
- **Other Heart Rhythm Abnormalities:** Numerous clinically significant heart rhythm abnormalities exist beyond those included in the current dataset used for model performance evaluation. Expanding the dataset to cover these additional conditions could enhance the model’s generalizability and clinical utility.
- **Testing at the Edge:** With continued advancements in hardware and code optimization, the novel KAN network can be deployed on-device for future testing.

### 8.2 Future Work

Future work should focus on addressing these limitations by:

- **Integration with Other Architectures:** Exploring the integration of KANs with other neural network architectures to create more robust and versatile models that could potentially perform better on certain abnormalities.
- **Code and Algorithm Optimization:** Enhancing the performance and reducing the computational complexity of KANs by optimizing their code and algorithms.
- **Deployment on Devices:** Deploying KANs on portable and wearable health monitoring systems, which requires further minimization of power requirements and ensuring sustainable and efficient operation in real-time applications.
- **Comprehensive Model:** KAN is a new model that still requires improvement. Future work will focus on developing a comprehensive ECG abnormality detector using more complex architectures and ensemble methods to enhance accuracy across all abnormality types.
- **Novel Training Integration:** The proposed efficient network may serve as a promising foundational model for on-device fine-tuning, facilitating model personalization while preserving data security through novel training and optimization mechanisms [42, 43]. This represents an emerging and intellectually stimulating research direction warranting further experimental investigation.

## 9 Conclusion

In this paper, we evaluate the performance of KAN and a shallow KAN-based network for ECG signal analysis, focusing on the detection of cardiac abnormalities, and compare them with traditional architectures such as MLPs and NCPs. Drawing inspiration from the Kolmogorov–Arnold representation theorem, KANs incorporate learnable activation functions, offering a potentially novel, efficient, and lightweight approach for edge applications. Our results highlight the promise of KANs for further optimization and potential deployment in realworld healthcare settings.

Our findings highlight the potential of KANs to enhance ECG analysis in cardiac healthcare, promising advancements in accuracy, interpretability, and efficiency. The results show that the KAN and CKAN model exhibits strong in-sample performance while maintaining good generalization in out-of-distribution scenarios. The model also shows robustness to incomplete data, which is common in real-world scenarios. When tested with single-lead ECG data instead of 12 leads, the model’s performance remains satisfactory, confirming its suitability for real-life applications.

The integration of learnable sparsity, a key feature of NCP neurons, into KAN neurons yielded surprisingly strong results. It not only enhanced the model’s performance and generalization but also facilitated the pruning of less important weights, making the model more efficient for TinyML applications.

While KANs have shown promising results, future research should focus on optimizing KANs for deployment on portable and wearable devices, ensuring they are efficient, robust, and capable of real-time data processing. These advancements could ultimately improve patient outcomes and more effective cardiac healthcare solutions.

## Data Availability

This paper uses the Chinese Physiological Signal Challenge 2018 (CPSC) dataset for analysis and experiments, which is openly available. The Telehealth Network of Minas Gerais (TNMG) dataset is not openly accessible. Access and permission to this data should be discussed with and granted by the custodian of the data.

## 10 Author Contribution Declaration

**Zhaojing Huang**: Led in conceptualization and methodology; led data curation, formal analysis, and investigation; contributed equally to software development, original draft writing, and review & editing; provided supporting roles in validation and visualization. **Jiashuo Cui**: Largely contributed to conceptualization and methodology; supported in validation and visualization; contributed to software development, original draft writing, and review & editing.

**Leping Yu**: Provided supporting contributions to original draft writing and review & editing.

**Luis Fernando Herbozo Contreras**: Provided supporting contributions to conceptualization and methodology.

**Girish Dwivedi**: Delivered clinical supervision and supported the review and refinement of the manuscript.

**Omid Kavehei**: Led supervision and provided supporting contributions to review & editing.

## 11 Acknowledgement

Zhaojing Huang acknowledges the support of the Australian Government’s Research Training Program (RTP).

## 12 Availability of Code

Data and relevant code for this research work are stored in GitHub: https://github.com/NeuroSyd/KAN-ECG and have been archived within the Zenodo repository: DOI: 10.5281/zenodo.13977105 [44]

## 13 Availability of Data

Two datasets are used in this paper, the Chinese Physiological Signal Challenge 2018 (CPSC) and the Telehealth Network of Minas Gerais (TNMG). The CPSC dataset is publicly available and can be accessed in the Challenge Data section at the following link: CPSC 2018. A permanent DOI for the dataset is accessible here: CPSC Dataset DOI [23]. The TNMG dataset is, however, not publicly available but it is accessible via requests to the custodian, who can be contacted through the corresponding author of Ref. [4].

## 14 Ethics Statement

Ethics, Consent to Participate, and Consent to Publish declarations: not applicable.

## 15 Funding Statement

Funding declaration: not applicable.

## 16 Conflict of Interest Statement

The authors declare no conflicts of interest.

## Notes

### Competing Interest Statement

The authors have declared no competing interest.

### Summary of Updates

Additional experiments and results included.

